# Pyrethroid resistance in *Anopheles gambiae* not associated with insecticide-treated mosquito net effectiveness across sub-Saharan Africa

**DOI:** 10.1101/2020.03.27.20045443

**Authors:** David A. Larsen, Rachael L. Church

**Affiliations:** Syracuse University Department of Public Health, Syracuse, New York, USA

## Abstract

**Background:** Pyrethroid resistance is a major concern for malaria vector control programs that predominantly rely on insecticide-treated mosquito nets (ITN). Contradictory results of the impact of resistance have been observed in field studies.

**Methods:** We combined continent-wide estimates of pyrethroid resistance in *Anopheles gambiae* from 2006-2017 with continent-wide survey data to assess the effect of increasing pyrethroid resistance on the effectiveness of ITNs to prevent malaria infections in sub-Saharan Africa. We utilized both a pooled-data approach and meta-regression of survey regions to assess how pyrethroid resistance affects the association between ITN ownership and malaria outcomes in children aged 6-59 months.

**Findings:** ITN ownership reduced the risk of malaria outcomes in both pooled and meta-regression approaches. In the pooled analysis, there was no observed interaction between ITN ownership and estimated level of pyrethroid resistance (Likelihood ratio [LR] test = 1.127 for the outcome of rapid diagnostic test confirmed malaria infection, p = 0.2885; LR test = 0.161 for the outcome of microscopy confirmed malaria infection, p = 0.161; LR test = 0.646 for the outcome of moderate or severe anemia, p = 0.4215). In the meta-regression approach the level of pyrethroid resistance did not explain any of the variance in subnational estimates of ITN effectiveness for any of the outcomes.

**Interpretation:** ITNs decreased risk of malaria outcomes independent of the levels of pyrethroid resistance in the malaria vector populations.

**Funding:** DAL did not receive funding and RC received a SOURCE grant from Syracuse University for this project.

## Introduction

Insecticide-treated mosquito nets (ITN) have been one of the most effective public health interventions of the 21^st^ century, preventing millions of deaths from the malaria parasite [1]. Previous to their wide-scale deployment, more than 1 million people died annually across sub-Saharan Africa from malaria infections [2], particularly the *Plasmodium falciparum* parasite, and the parasite caused a further 500 million cases. Broad scale-up of ITNs and across the continent in the early and mid-2000’s [3] contributed to this annual burden being cut in half by 2015 [4]. The 2018 World Malaria Report found that all species of malaria were responsible for 435,000 global deaths and 219 million global cases in 2017 [5], a huge reduction from estimates earlier this century. It is worrisome, however, that the 2018 World Malaria Report found the first annual increase in global malaria cases since 2005.

Mosquito nets provide a personal barrier to prevent infective bites from malaria vectors that predominantly bite at night [6]. An untreated mosquito net will protect the person sleeping beneath from an infective bite of an infected mosquito. However, that infective malaria vector will likely seek a blood meal elsewhere and thus continue malaria transmission. When the mosquito net is treated with insecticide and the mosquito is susceptible, contact with the insecticide kills the mosquito and ends that mosquito’s capacity to continue transmitting malaria [7]. The subsequent effect of ITNs on malaria transmission is profound [8]. When a high number of ITNs are used within a community, a mass killing effect from the insecticide provides a community-level protection, sufficiently affecting the malaria vector populations such that people who do not have an ITN are protected in similar ways to those that do have an ITN [9,10].

Currently, pyrethroids are the only class of insecticides used on ITNs [11], although combination nets treated with multiple insecticides are in various stages of testing [12–14]. Pyrethroids target the central nervous system to kill mosquitoes and reduce the transmission of malaria by incapacitating the vector [15]. Fears about malaria vectors developing pyrethroid resistance were raised at the inception of ITNs [15], and early studies demonstrated widespread reports of resistance [16,17]. Although alarming, it remains to be seen just how pyrethroid resistance will affect malaria control and malaria trends [18–20].

Linking pyrethroid resistance in malaria vectors to ITN effectiveness to prevent malaria infections and reduce malaria transmission has been more challenging than documenting the spread of resistance. Some studies suggest that in areas with pyrethroid resistance, ITNs are less effective [21,22]. Others find that ITNs are still associated with reduced risk of malaria despite high levels of pyrethroid resistance [23–26]. These studies generally struggle with four separate issues. First, due to the inability to randomly allocate ITN access, these studies suffer from selection bias wherein households with ITN access are more likely to be predisposed to lower risk of malaria transmission than households without ITN access independent of the effect of ITNs [27]. Second, some of these studies do not actually include estimates of ITN effectiveness, and the studies that do utilize ITN use as their measure of effectiveness. ITN use may not be the best measure to understand the impact of pyrethroid resistance on ITN effectiveness. Even if malaria vectors are resistant to the insecticide in the ITN, the mosquito netting still provides a physical barrier and consistent use will decrease mosquito exposure and thus malaria risk [28]. This is evident where in areas of pyrethroid resistance ITNs with holes lose their effectiveness [29]. Third, each of the studies is confined to a particular study site. Investigating phenomena at particular study sites limits the variability of resistance available, as the broad spectrum of resistance prevalence is not incorporated within the study. And fourth, the studies utilize an overly simplistic assessment of insecticide resistance, typically with dichotomized low resistance and high resistance areas that prevent an examination of a dose-response relationship between ITN effectiveness and pyrethroid resistance.

Herein we attempt to address these four challenges and better understand how widespread pyrethroid resistance is affecting the ability of ITNs to reduce malaria transmission. We leverage nationally representative surveys across sub-Saharan Africa and recently published near continent-wide estimates of pyrethroid resistance of *Anopheles gambiae* in sub-Saharan Africa[30] to address the issues of geographic limitation, a limited resistance spectrum, and oversimplification in the statistical measure of insecticide resistance. We further utilize exact matching to minimize the influence of selection bias, and a measure of ITN ownership rather than ITN use to reduce the residual confounding of the effect of mosquito netting as a barrier.

## Methods

### Search strategy and selection criteria

We utilized data from nationally representative Demographic and Health Surveys (DHS) to assess the relationship between insecticide resistance and the effectiveness of ITNs. All DHS surveys conducted in sub-Saharan Africa between 2006-2017 that were publicly available as of January 15, 2020 were considered for inclusion in the analyses with the following criteria: the survey contained information on an outcome (malaria infection status in children either as measured through rapid-diagnostic test or through microscopy, or anemia status as measured through a hemocue rapid hemoglobin assessment), and the survey contained information on household ITN ownership. We further limited these surveys to those that were conducted in areas with estimated prevalence of pyrethroid resistance [30]. The DHS program is funded by USAID and assists lower income countries in conducting nationally-representative surveys primarily aimed at measuring trends in child mortality and women’s fertility with statistical power at the regional (sub-national) level. Additionally, these surveys gather a host of information on numerous factors associated with health, including malaria indicators when the survey is conducted in a malaria-endemic country.

### Outcome measures

We considered three separate outcome measures in children aged 7-49 months: confirmation of a malaria infection through rapid diagnostic test, confirmation of a malaria infection through microscopy, or hemoglobin levels < 10 g/dl (moderate to severe anemia). Anemia is an historical indicator of malaria transmission, and is also known to be associated with effective malaria control such as ITNs [31].

### ITN ownership measure

We defined the primary exposure of ITN ownership as the child living in a household that owned at least one ITN.

### Exact matching of observations

Selection bias presents the largest challenge to validity in assessing ITN effectiveness in observational studies. We followed an exact matching approach similar to the analysis of ITN effectiveness conducted by Lim et al. to minimize this selection bias [32], wherein we matched households on the probability of owning an ITN. Specifically, we matched households within survey datasets on the following covariate pattern: region of the country as per the DHS, wealth categorized as rich or poor, mother’s education categorized as any or none, and the house being in an urban or rural location. Matching was done using the MatchIt package [33,34], in R version 3.6.2 [35].

### Statistical analysis

We utilized two separate approaches to assess the influence of pyrethroid resistance on the effectiveness of insecticide-treated mosquito nets.

#### Pooled analysis approach

We combined all survey data meeting inclusion criteria and exactly matched into a single dataset. We extracted the estimates of resistance, seasonality of malaria transmission from the Mapping Malaria Risk in Africa (MARA) maps [36], estimates of the *Plasmodium falciparum* parasite prevalence rate in children aged 2-10 (*Pf*PR) from the Malaria Atlas Project [37], and the relative abundance of the *Anopheles gambiae* vector [38] to the cluster geocoordinates using the Raster package [39] in R version 3.6.2 [35]. *Pf*PR estimates were only available for the years 2000-2015 from the MalariaAtlas package, and so we assigned surveys conducted 2016-2017 *Pf*PR levels of 2015 (n= 14 surveys).

We then estimated the relationship between ITN effectiveness and the prevalence of resistance using an interaction term predicting the outcome using a logistic regression approach with the matched group as a random intercept (Equation 1). We adjusted analyses for child’s age categorized as years (<1, 1, 2, 3, 4), household wealth quintile, mother’s education (none, some primary, completed primary or higher), whether the data were collected during the transmission season or not, household location (urban or rural), and *Pf*PR levels (continuous). The three separate outcomes were modeled in the same fashion., one for the outcome of RDT-confirmed malaria infection and the other for the outcome of microscopy-confirmed malaria infection. We conducted this analysis for all observations in the dataset, and then limited the analysis to those observations where the relative abundance of *An. gambiae* was > 0.5. The analyses were conducted using Stata version 15.1.

*Equation 1: Representation of pooled analysis wherein π*_*ijklm*_ *is a dichotomous outcome for child i in household j in cluster k in survey l in matched group m, ITN*_*j*_ *is whether the household has access to any ITN or not, Res*_*k*_ *is the level of pyrethroid resistance access in the community, C*_*ijk*_ *is a vector of child characteristics, H*_*j*_ *is a vector of household characteristics, S*_*k*_ *is a vector of cluster characteristics, and ζ*_*m*_ *is a random intercept for matched group m that is assumed to be normally distributed with a mean of zero*.

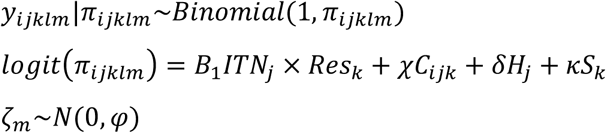

#### Meta-regression approach

We parsed all survey data meeting inclusion criteria to DHS region level. We extracted the estimates of pyrethroid resistance, the *Plasmodium falciparum* parasite prevalence rate in children aged 2-10 (*Pf*PR) from the Malaria Atlas Project [37], and the relative abundance of the *Anopheles gambiae* vector[38] to the DHS region level using the Raster package [39] in R version 3.6.2 [35]. *Pf*PR estimates were only available for the years 2000-2015 from the MalariaAtlas package, and so we assigned surveys conducted 2016-2017 *Pf*PR levels of 2015 (n= 14 surveys).

We then estimated the effectiveness of ITNs for each DHS region as described previously using equation 1. We conducted a meta-regression of ITN effectiveness within DHS regions using the metafor package [40] in R version 3.6.2 [35] with the prevalence of pyrethroid resistance at the region level serving as the predictor. We also tested for interactions between pyrethroid resistance and both *Pf*PR as well as the abundance of *Anopheles gambiae*.

### Role of the funding source

DAL was unfunded for this work. RC received a SOURCE grant from Syracuse University. The sponsors had no role in the study design; collection, analysis, and interpretation of the data; in the writing of the report; and in the decision to submit the paper for publication. DAL had full access to all the data in the study and had final responsibility for the decision to submit for publication.

## Results

### Data available

We identified 92 survey datasets conducted in sub-Saharan Africa between 2006-2017 publicly available as of January 15, 2020 with information on ITN ownership, malaria infection in children, or child’s anemia status. Of these 92 surveys, 30 were excluded from any analysis due to the following reasons: 12 surveys contained no information on malaria infection or anemia status, 5 surveys contained no information on household ITN ownership, and 8 surveys could not be matched to resistance estimates (the surveys were conducted in Angola, Sao Tome and Principe, or Madagascar where no resistance estimates were available). A further seven surveys had no cluster geocoordinates and so were excluded from the pooled analysis but included in the meta-regression approach. Figure 1 shows the geographical distribution of 67 surveys included in the analyses.

**Figure 1:**
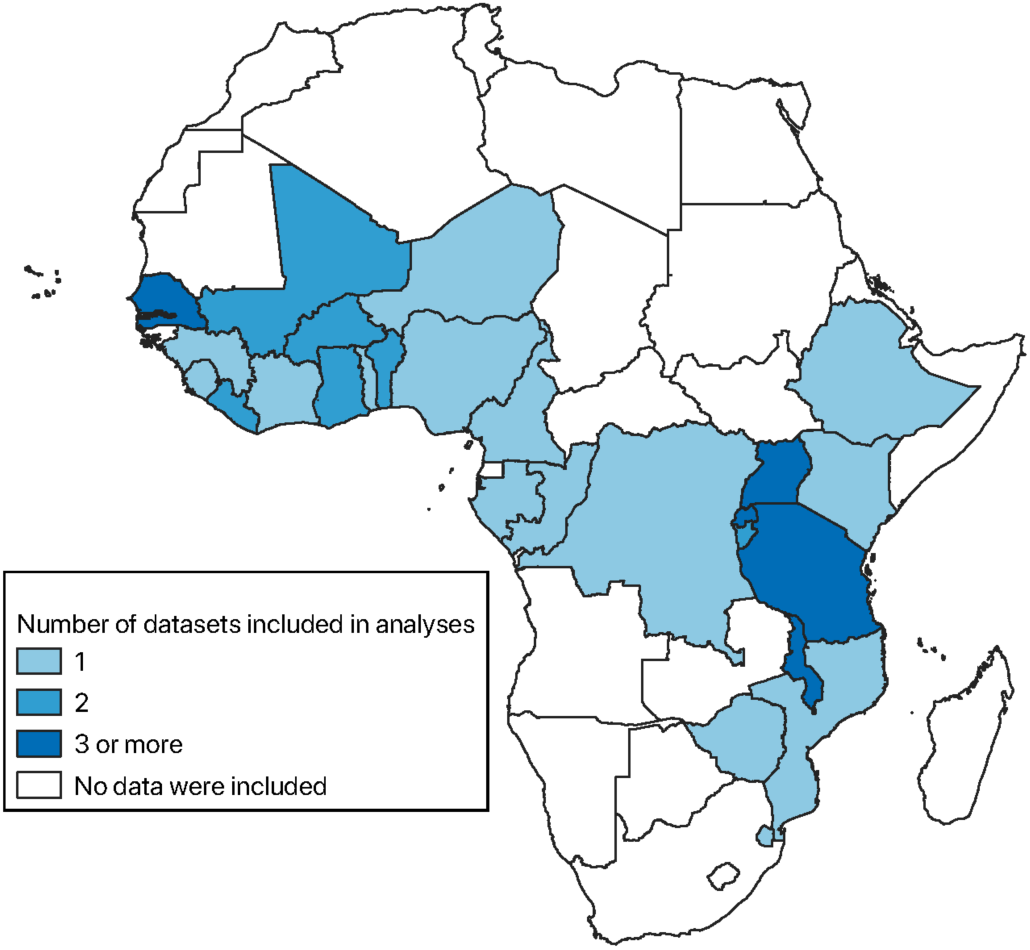
Geographic distribution of data included in the analyses.

### Variation of pyrethroid resistance

At the survey cluster level, estimates of pyrethroid resistance within this dataset ranged from 10.5% to 99.9% with a median level of 89.1% (Figure 2). Pyrethroid resistance was associated with decreased levels of *Pf*PR within survey clusters (rho = -0.3061, n = 18,927 clusters) and decreased relative abundance of *An. gambiae* (rho = -0.2321, n = 19,588 clusters).

**Figure 2:**
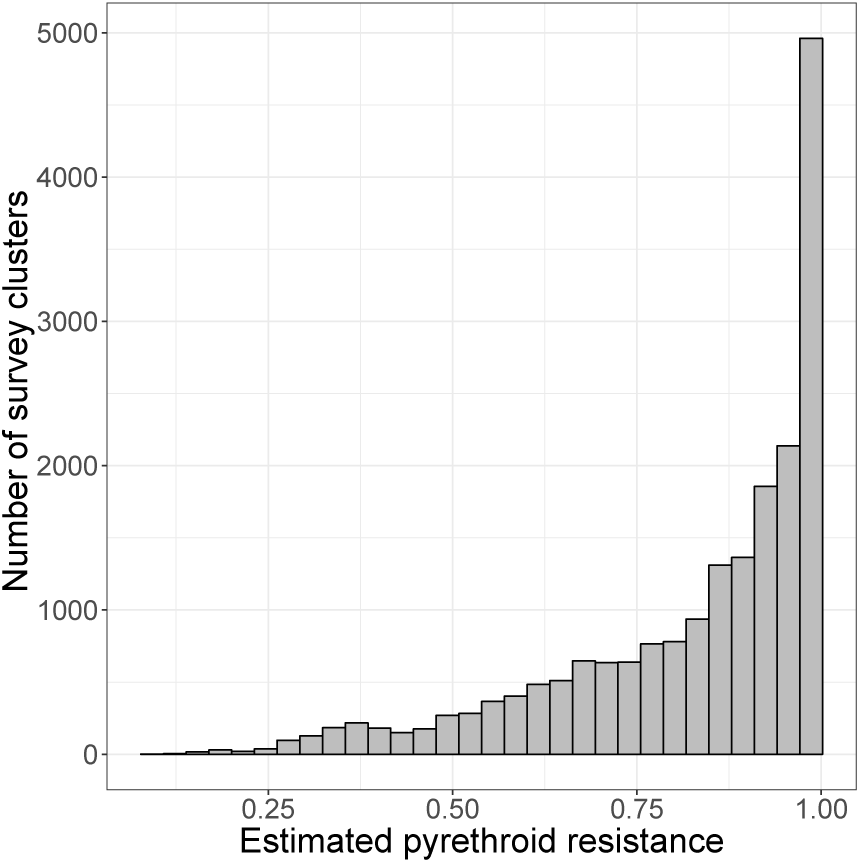
Distribution of estimated pyrethroid resistance across datasets included in the analyses.

### Effect of pyrethroid resistance on association between ITNs and malaria outcomes – pooled analysis approach

After exact matching and adjusting for known predictors but without accounting for resistance, ownership of an ITN was associated with a reduction in the risk of a malaria infection as well as having moderate or severe anemia (Table 1). After accounting for resistance, there was no evidence of an interaction between ITN ownership and pyrethroid resistance levels for any of the outcomes (Table 2). ITN ownership was still associated with a reduction in the risk of a malaria infection as well as having moderate or severe anemia. Limiting the analysis to areas with >50% *An. gambiae* did not change the results.

**Table 1:** Association between ITN ownership and various outcomes without taking pyrethroid resistance into account. Children were matched on probability of ITN ownership. Models were adjusted for urban/rural, child’s age, wealth quintile, mother’s education, PfPR, malaria transmission season, and survey dataset.

| Outcome | Adjusted odds ratio (95% CI) of ITN ownership | P-value | N children (n matched groups) |
| --- | --- | --- | --- |
| **RDT positivity** | 0.90 (0.87 – 0.93) | < 0.001 | 137,943 (4,493) |
| **Microscopy positivity** | 0.88 (0.84 – 0.91) | < 0.001 | 116,556 (3,643) |
| **Moderate or severe anemia** | 0.95 (0.92 – 0.97) | < 0.001 | 163,332 (5,486) |

**Table 2:** Likelihood ratio test for including an interaction term for pyrethroid resistance and ITN ownership. Children were matched on probability of ITN ownership. Models were adjusted for urban/rural, child’s age, wealth quintile, mother’s education, PfPR, malaria transmission season, and survey dataset.

| **Outcome** | **Likelihood ratio test for interaction between pyrethroid resistance and ITN ownership** | **P-value** | **N children (n matched groups)** |
| --- | --- | --- | --- |
| **RDT positivity** | 1.127 | 0.2885 | 137,943 (4,493) |
| **Microscopy positivity** | 0.161 | 0.6880 | 116,556 (3,643) |
| **Moderate or severe anemia** | 0.646 | 0.4215 | 163,332 (5,486) |

### Effect of pyrethroid resistance on association between ITNs and malaria outcomes – meta-regression approach

Within-DHS region specific estimates of the effectiveness of ITNs were available for 349 regions for the outcome of RDT-confirmed malaria infection, 243 regions for the outcome of microscopy-confirmed malaria infection, and 611 regions for the outcome of moderate or severe anemia. In unadjusted meta-analyses, ITN ownership was associated with a reduced risk of RDT-confirmed malaria infection and microscopy-confirmed malaria infection, but not moderate or severe anemia (Table 3). Pyrethroid resistance did not explain any of the regional variance in the effectiveness of ITN ownership against the outcomes (Figure 3), nor did any interaction between pyrethroid resistance and either *Pf*PR or the abundance of *An. gambiae*.

**Table 3:** Results from unadjusted meta-analyses of ITN ownership on the outcomes. DHS data were parsed to region. Children were matched on probability of ITN ownership. Region-specific models were adjusted for urban/rural, child’s age, wealth quintile, and mother’s education.

| **Outcome** | **Odds ratio (95% Confidence Interval)** | **P-value** | **N DHS regions** | **I-squared** |
| --- | --- | --- | --- | --- |
| **RDT positivity** | 0.888 (0.833 – 0.947) | < 0.001 | 349 | 0.00% |
| **Microscopy positivity** | 0.857 (0.795 – 0.925) | < 0.001 | 243 | 0.00% |
| **Moderate or severe anemia** | 0.965 (0.921 – 1.011) | 0.1330 | 611 | 0.00% |

**Figure 3:**
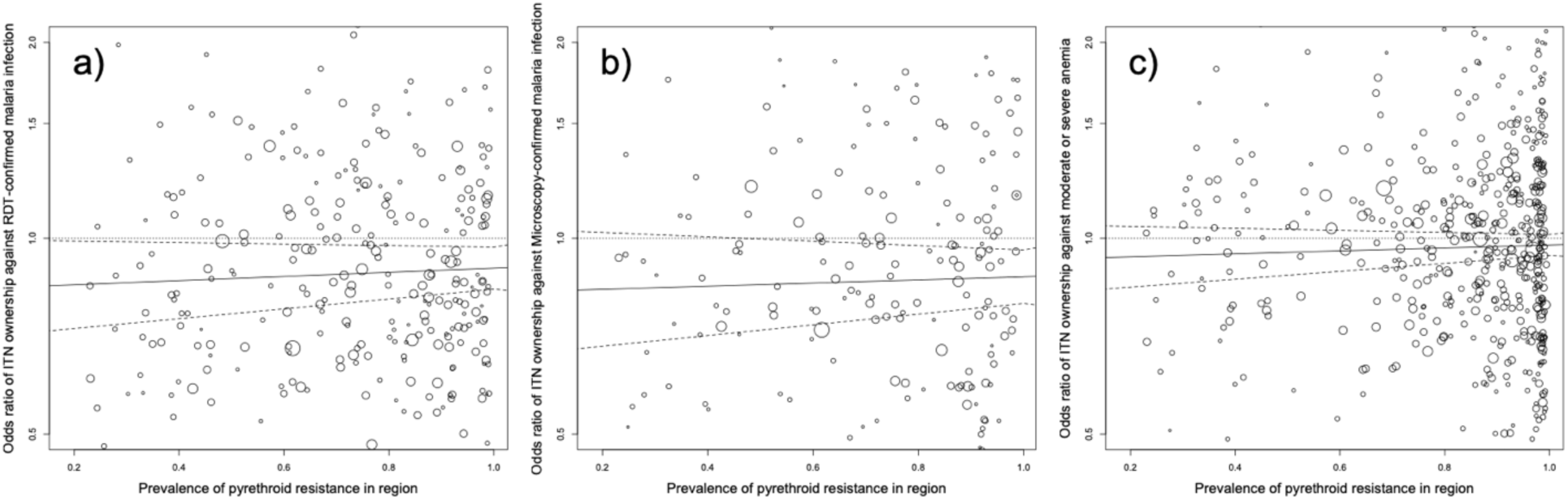
Bubble plots showing the relationship between pyrethroid resistance at the region level and effectiveness of ITN ownership against a) RDT-confirmed malaria infection, b) microscopy-confirmed malaria infection, and c) moderate or severe anemia.

## Discussion

### Key results

These results suggest that the effectiveness of ITNs is not affected by the prevalence of pyrethroid resistance across sub-Saharan Africa. ITN ownership was associated with a reduced risk of a malaria infection (either RDT- or microscopy-confirmed) as well as a reduced risk of moderate or severe anemia. These reductions were modest, just more than 10% for the malaria infection and 5% for moderate severe anemia. Importantly, however, pyrethroid resistance did not modify these reductions in any way. There was also agreement across both the pooled and meta-regression approaches utilized herein.

### Limitations

Selection bias is present in the data, and occurs either as houses owning ITNs are predisposed to less malaria transmission independent of ITN ownership or ITN distribution programs are targeted toward areas of higher malaria transmission and therefore children in houses owning ITNs are predisposed to more malaria transmission independent of ITN ownership. We attempted to mitigate the effects of this selection bias on the analysis using exact matching, as we and others have done in previous analyses of ITN ownership using these data [9,32]. It is possible that some residual bias is still present, and if so we would expect that bias to overinflate the effect of ITN ownership. However, should there be a decrease in ITN effectiveness with increasing pyrethroid resistance we would still expect to see that effect given the limitations in the data.

Pyrethroid resistance estimates for these data were quite high, with a median of 89%. Unfortunately, with such high levels of pyrethroid resistance spread across the continent we could not examine the effectiveness of ITNs across the entire range from 0-100% resistance. Given the large volume of data we included in our analyses we still had sufficient power to determine that ITN ownership reduces the risk of malaria infection even when estimated pyrethroid resistance levels approach 100% of the vector population.

### Interpretation

These results are positive news for malaria control program. ITNs remain the primary vector control strategy throughout sub-Saharan Africa, and they appear to be effective even at high levels of pyrethroid resistance. Others have described in detail this resistance paradox, where in laboratory experiments pyrethroid resistance confers protection for malaria vectors but in epidemiological studies vector control confers protection against malaria infection [41]. In general two separate hypotheses are present as to how ITNs maintain effectiveness in the face of widespread resistance. The first hypothesis suggests that the same genes that confer pyrethroid resistance alter the ability of the vector to transmit the parasite. A fitness cost to pyrethroid resistance has been documented in the *An. gambiae*, as evidenced by studies showing that non-resistant mosquitoes outcompete resistant mosquitoes [42]. The second hypothesis suggests that the insecticide still functions, just with a different type of effect. One example of this second hypothesis is that pyrethroid-resistant mosquitoes lose their irritancy to the insecticide, thus allowing for increased contact and a higher dose of the insecticide than typical mosquitoes [43]. This increased dose then leads to the mosquito’s death. Another example of this second hypothesis is that a dose of pyrethroids results in delayed mortality of pyrethroid-resistant mosquitoes [44], perhaps due to impaired flight or behavioral effects of the insecticide [45]. Whatever the reason, ITNs still appear to be effectively reducing malaria transmission even when pyrethroid resistance approaches 100% in vector populations.

## Conclusion

Insecticide resistance is a chief concern for malaria control programs. These results confirm the notion that pyrethroid-based ITNs are still working to prevent malaria despite high levels of pyrethroid resistance observed in malaria vector populations.

## Data Availability

All data used in this study are publicly available from the Demographic and Health Survey Project or through references.

## Declaration of interests

The authors have no conflict of interests.

## Notes

### Competing Interest Statement

The authors have declared no competing interest.

### Funding Statement

RLC was supported by a student research award from Syracuse University

## References

1. Eisele TP, Larsen DA, Walker N, Cibulskis RE, Yukich JO, Zikusooka CM, et al. Estimates of child deaths prevented from malaria prevention scale-up in Africa 2001-2010. Malar J. 2012;11: 93. doi:10.1186/1475-2875-11-93

2. Murray CJL, Rosenfeld LC, Lim SS, Andrews KG, Foreman KJ, Haring D, et al. Global malaria mortality between 1980 and 2010: a systematic analysis. Lancet. Institute for Health Metrics and Evaluation, University of Washington, Seattle, WA 98121, USA.: Elsevier; 2012;379: 413–431. Available: http://eutils.ncbi.nlm.nih.gov/entrez/eutils/elink.fcgi?dbfrom=pubmed&id=22305225&retmode=ref&cmd=prlinks

3. Bhatt S, Weiss DJ, Mappin B, Dalrymple U, Cameron E, Bisanzio D, et al. Coverage and system efficiencies of insecticide-treated nets in Africa from 2000 to 2017. Elife. 2015;4: e09672. doi:10.7554/eLife.09672

4. Bhatt S, Weiss DJ, Cameron E, Bisanzio D, Mappin B, Dalrymple U, et al. The effect of malaria control on Plasmodium falciparum in Africa between 2000 and 2015. Nature. 2015;526: 207–11. doi:10.1038/nature15535

5. World Health Organisation. World Malaria Report. World Malaria Report. 2018. doi: ISBN 978 92 4 156483 0

6. Rund SSC, O’Donnell AJ, Gentile JE, Reece SE. Daily rhythms in mosquitoes and their consequences for malaria transmission. Insects. 2016;7: 1–20. doi:10.3390/insects7020014

7. Sutcliffe JF, Yin S. Behavioural responses of females of two anopheline mosquito species to human-occupied, insecticide-treated and untreated bed nets. Malar J. 2014;13: 294. doi:10.1186/1475-2875-13-294

8. Lengeler C. Insecticide-treated bed nets and curtains for preventing malaria. Cochrane Database Syst Rev. 2004; CD000363.

9. Larsen DA, Hutchinson P, Bennett A, Yukich J, Anglewicz P, Keating J, et al. Community coverage with insecticide-treated mosquito nets and observed associations with all-cause child mortality and malaria parasite infections. Am J Trop Med Hyg. 2014;91: 950–958. doi:10.4269/ajtmh.14-0318

10. Hawley WA, Phillips-Howard PA, Ter Kuile FO, Terlouw DJ, Vulule JM, Ombok M, et al. Community-wide effects of permethrin-treated bed nets on child mortality and malaria morbidity in western Kenya. Am J Trop Med Hyg. 2003;68: 121–127.

11. Ranson H, Lissenden N. Insecticide Resistance in African Anopheles Mosquitoes: A Worsening Situation that Needs Urgent Action to Maintain Malaria Control. Trends in Parasitology. 2016. pp. 187–196. doi:10.1016/j.pt.2015.11.010

12. Cisse MBM, Sangare D, Oxborough RM, Dicko A, Dengela D, Sadou A, et al. A village level cluster-randomized entomological evaluation of combination long-lasting insecticidal nets containing pyrethroid plus PBO synergist in Southern Mali. Malar J. 2017;16: 477. doi:10.1186/s12936-017-2124-1

13. Gleave K, Lissenden N, Richardson M, Choi L, Ranson H. Piperonyl butoxide (PBO) combined with pyrethroids in insecticide-treated nets to prevent malaria in Africa. Cochrane Database Syst Rev. 2018;2018. doi:10.1002/14651858.CD012776.pub2

14. Tiono AB, Ouédraogo A, Ouattara D, Bougouma EC, Coulibaly S, Diarra A, et al. Efficacy of Olyset Duo, a bednet containing pyriproxyfen and permethrin, versus a permethrin-only net against clinical malaria in an area with highly pyrethroid-resistant vectors in rural Burkina Faso: a cluster-randomised controlled trial. Lancet. 2018;0. doi:10.1016/S0140-6736(18)31711-2

15. Hemingway J, Ranson H. Insecticide Resistance in Insect Vectors of Human Disease. Annu Rev Entomol. 2000;45: 371–391. doi:10.1146/annurev.ento.45.1.371

16. Santolamazza F, Calzetta M, Etang J, Barrese E, Dia I, Caccone A, et al. Distribution of knock-down resistance mutations in Anopheles gambiae molecular forms in west and west-central Africa. Malar J. 2008;7: 1–8. doi:10.1186/1475-2875-7-74

17. Ranson H, N’guessan R, Lines J, Moiroux N, Nkuni Z, Corbel V. Pyrethroid resistance in African anopheline mosquitoes: what are the implications for malaria control? Trends Parasitol. Vector Group, Liverpool School of Tropical Medicine, Pembroke Place, Liverpool, UK, L3 5QA.; 2011;27: 91–98. Available: http://eutils.ncbi.nlm.nih.gov/entrez/eutils/elink.fcgi?dbfrom=pubmed&id=20843745&retmode=ref&cmd=prlinks

18. Hemingway J, Ranson H, Magill A, Kolaczinski J, Fornadel C, Gimnig J, et al. Averting a malaria disaster: Will insecticide resistance derail malaria control? Lancet. 2016;387: 1785–1788. doi:10.1016/S0140-6736(15)00417-1

19. Kelly-Hope L, Ranson H, Hemingway J. Lessons from the past: managing insecticide resistance in malaria control and eradication programmes. Lancet Infect Dis. 2008;8: 387–389. doi:10.1016/S1473-3099(08)70045-8

20. Thomas MB, Read AF. The threat (or not) of insecticide resistance for malaria control. Proc Natl Acad Sci. 2016;113: 8900–8902. doi:10.1073/pnas.1609889113

21. Opondo KO, Weetman D, Jawara M, Diatta M, Fofana A, Crombe F, et al. Does insecticide resistance contribute to heterogeneities in malaria transmission in the Gambia? Malar J. BioMed Central; 2016;15: 1–10. doi:10.1186/s12936-016-1203-z

22. Mathanga DP, Mwandama DA, Bauleni A, Chisaka J, Shah MP, Landman KZ, et al. The effectiveness of long-lasting, insecticide-treated nets in a setting of pyrethroid resistance: a case-control study among febrile children 6 to 59 months of age in Machinga District, Malawi. Malar J. BioMed Central; 2015;14: 1–8. doi:10.1186/s12936-015-0961-3

23. Kleinschmidt I, Bradley J, Knox TB, Mnzava AP, Kafy HT, Mbogo C, et al. Implications of insecticide resistance for malaria vector control with long-lasting insecticidal nets: a WHO-coordinated, prospective, international, observational cohort study. Lancet Infect Dis. 2018;18: 640–649. doi:10.1016/S1473-3099(18)30172-5

24. Tokponnon FT, Sissinto Y, Ogouyémi AH, Adéothy AA, Adechoubou A, Houansou T, et al. Implications of insecticide resistance for malaria vector control with long-lasting insecticidal nets: Evidence from health facility data from Benin. Malar J. BioMed Central; 2019;18: 1–9. doi:10.1186/s12936-019-2656-7

25. Bradley J, Ogouyèmi-Hounto A, Cornélie S, Fassinou J, De Tove YSS, Adéothy AA, et al. Insecticide-treated nets provide protection against malaria to children in an area of insecticide resistance in Southern Benin. Malar J. BioMed Central; 2017;16: 1–5. doi:10.1186/s12936-017-1873-1

26. Eric Ochomo, 1, Mercy Chahilu, 1, Jackie Cook, Teresa Kinyari, Nabie M. Bayoh PW, Luna Kamau, Aggrey Osangale, Maurice Ombok, Kiambo Njagi EM, et al. Insecticide-Treated Nets and Protection against Insecticide-Resistant Malaria Vectors in Western Kenya. Emerg Infect Dis. 2017;23: 158–764. doi:10.3201/eid2305.161315

27. Tusting LS, Bottomley C, Gibson H, Kleinschmidt I, Tatem AJ, Lindsay SW, et al. Housing Improvements and Malaria Risk in Sub-Saharan Africa: A Multi-Country Analysis of Survey Data. PLoS Med. 2017;14: 1–15. doi:10.1371/journal.pmed.1002234

28. Clarke SE, Bøgh C, Brown RC, Pinder M, Walraven GELL, Lindsay SW, et al. Do untreated bednets protect against malaria? Trans R Soc Trop Med Hyg. 2001;95: 457–462. doi:10.1016/S0035-9203(01)90001-X

29. Ochomo EO, Bayoh NM, Walker ED, Abongo BO, Ombok MO, Ouma C, et al. The efficacy of long-lasting nets with declining physical integrity may be compromised in areas with high levels of pyrethroid resistance. Malar J. 2013;12: 1–10. doi:10.1186/1475-2875-12-368

30. Hancock P, Hendricks C, Tangena J-A, Gibson H, Hemingway J, Coleman M, et al. Mapping trends in insecticide resistance phenotypes in African malaria vectors. bioRxiv. 2020;

31. Korenromp EL, Armstrong-Schellenberg JRM, Williams BG, Nahlen BL, Snow RW. Impact of malaria control on childhood anaemia in Africa -- a quantitative review. Trop Med Int Health. World Health Organization, Roll Back Malaria, Geneva, Switzerland.; 2004;9: 1050–1065.

32. Lim SS, Fullman N, Stokes A, Ravishankar N, Masiye F, Murray CJL, et al. Net Benefits: A Multicountry Analysis of Observational Data Examining Associations between Insecticide-Treated Mosquito Nets and Health Outcomes. PLoS Med. 2011;8: e1001091. Available: http://www.mara.org/

33. Ho D, Imai K, King G, Stuart EA. MatchIt: Nonparametric preprocessing for parametric causal inference. Citeseer. 2007; Available: http://citeseerx.ist.psu.edu/viewdoc/download?doi=10.1.1.138.698&rep=rep1&type=pdf

34. Ho DE, Imai K, King G, Stuart EA. Matching as Nonparametric Preprocessing for Reducing Model Dependence in Parametric Causal Inference. Polit Anal. 2007;15: 199–236. Available: http://gking.harvard.edu/files/abs/matchp-abs.shtml

35. R Core Development Team. R: A Language and Environment for Statistical Computing. http://www.R-project.org/. R Foundation for Statistical Computing; 2010;

36. Craig MH, Snow RW, le Sueur D. A climate-based distribution model of malaria transmission in sub-Saharan Africa. Parasitol today (Personal ed). National Malaria Research Programme, South African Medical Research Council, PO Box 17120, Congella, Durban 4013, South Africa.; 1999;15: 105–111. Available: http://eutils.ncbi.nlm.nih.gov/entrez/eutils/elink.fcgi?dbfrom=pubmed&id=10322323&retmode=ref&cmd=prlinks

37. Gething PW, Patil AP, Smith DL, Guerra CA, Elyazar IRF, Johnston GL, et al. A new world malaria map: Plasmodium falciparum endemicity in 2010. Malar J. Spatial Ecology and Epidemiology Group, Tinbergen Building, Department of Zoology, University of Oxford, South Parks Road, Oxford, UK.: Malaria Journal; 2011;10: 378. Available: http://eutils.ncbi.nlm.nih.gov/entrez/eutils/elink.fcgi?dbfrom=pubmed&id=22185615&retmode=ref&cmd=prlinks

38. Sinka ME, Bangs MJ, Manguin S, Coetzee M, Mbogo CM, Patil AP, et al. The dominant Anopheles vectors of human malaria in Africa, Europe and the Middle East: occurrence data, distribution maps and bionomic précis. Parasit Vectors. 2011;4: 89. doi:10.1186/1756-3305-4-89

39. Hijmans RJ. Introduction to the’raster’package (version 2.0-08). 2012; Available: http://probability.ca/cran/web/packages/raster/vignettes/Raster.pdf

40. Viechtbauer W. Conducting meta-analyses in R with the metafor package. J Stat Softw. 2010;36. Available: http://www.jstatsoft.org/v36/i03/paper

41. Alout H, Labbé P, Chandre F, Cohuet A. Malaria Vector Control Still Matters despite Insecticide Resistance. Trends Parasitol. Elsevier Ltd; 2017;33: 610–618. doi:10.1016/j.pt.2017.04.006

42. Lynd A, Weetman D, Barbosa S, Egyir Yawson A, Mitchell S, Pinto J, et al. Field, genetic, and modeling approaches show strong positive selection acting upon an insecticide resistance mutation in anopheles gambiae s.s. Mol Biol Evol. 2010;27: 1117–1125. doi:10.1093/molbev/msq002

43. Chandre F, Darriet F, Duchon S, Finot L, Manguin S, Carnevale P, et al. Modifications of pyrethroid effects associated with kdr mutation in Anophelos gambiae. Med Vet Entomol. 2000;14: 81–88. doi:10.1046/j.1365-2915.2000.00212.x

44. Viana M, Hughes A, Matthiopoulos J, Ranson H, Ferguson HM. Delayed mortality effects cut the malaria transmission potential of insecticide-resistant mosquitoes. Proc Natl Acad Sci. 2016;113: 8975–8980. doi:10.1073/pnas.1603431113

45. Cohnstaedt LW, Allan SA. Effects of sublethal pyrethroid exposure on the host-seeking behavior of female mosquitoes. J Vector Ecol. 2011;36: 395–403. doi:10.1111/j.1948-7134.2011.00180.x

